# Structural Covariance Analysis of Altered Brain Development in Neonates with Congenital Heart Disease After Surgery

**DOI:** 10.64898/2026.04.06.26350234

**Authors:** Mirthe E.M. van der Meijden, Barat Gal-Er, Benjamin Clayden, Siân Wilson, Daniel Cromb, Andrew Chew, Alexia Egloff, Kuberan Pushparajah, John Simpson, Joseph V. Hajnal, A. David Edwards, Mary Rutherford, Jonathan O’Muircheartaigh, Serena J. Counsell, Alexandra F. Bonthrone

**Author notes:** **Corresponding author:** Dr Alexandra F. Bonthrone. Joint senior authors (Serena J. Counsell, Alexandra F. Bonthrone).

## Abstract

**Background:** Brain development is altered in neonates with congenital heart disease (CHD). However, development in the perioperative period remains incompletely understood.

**Purpose:** This study used Structural Covariance Component (SCC) analysis to identify brain regions showing spatial patterns of coordinated expansion and contraction that differ between neonates with CHD after cardiac intervention and healthy controls, as well as pre-to postoperative changes and effects of perioperative risk factors.

**Study type:** Prospective.

**Population:** The cohort included 41 neonates with CHD who underwent cardiac surgery or catheterization and 359 healthy neonates.

**Field strength and sequence:** 3 Tesla T2-weighted turbo-spin-echo sequence.

**Assessment:** Brain MRI were motion-corrected and reconstructed using an established neonatal algorithm. Jacobian determinants calculated from non-linear registration of MRI to a neonatal template were input into an Independent Component Analysis to identify SCCs (N=40). SCC weightings were extracted, reflecting the degree to which the pattern of covariance is expressed in each neonate.

**Statistical tests:** Postoperative SCC weightings were compared to healthy neonates using a general linear model or robust regression. Pre- and postoperative SCC weightings were compared using a linear mixed effect model. Pre- to postoperative differences were calculated and associations with age at surgery, cardiopulmonary bypass duration, and postoperative paediatric intensive care unit stay were assessed using partial spearman’s rank correlation. Analyses were adjusted for covariates and corrected for multiple comparisons using False Discovery Rate.

**Results:** 16/40 SCCs showed significant differences between neonates with CHD after surgery and controls, including white matter, cortical- and deep grey matter, brainstem, and CSF regions, with seven also showing significant perioperative change. A further nine SCCs only showed significant perioperative change. Perioperative risk factors were not associated with perioperative change.

**Data conclusion:** This data-driven approach highlights region-specific postoperative alterations and perioperative changes in brain morphology of neonates with CHD. **Evidence level.** 1.

**Technical Efficacy:** Stage 3.

## Introduction

Congenital heart disease (CHD) is the most prevalent congenital defect, comprising structural abnormalities of the heart and great vessels (1). Neonates with CHD are at increased risk of altered brain development and neurodevelopmental impairments (2–4), which remain common despite substantial improvements in surgical and perioperative care (5). Characterising altered brain development in the perioperative period is essential for informing future approaches to improve neurodevelopmental outcomes in CHD.

MRI studies report that altered brain development in CHD originates *in utero*, where abnormal development of cardiac structures leads to suboptimal cerebral oxygen delivery (6–9), and persists into the preoperative neonatal period (10–12). Following neonatal cardiac surgery, smaller brain volumes (13), reduced brain growth (14), and new white matter injury (15) have been reported, emphasizing that the perioperative period may play a critical role in brain development, potentially compounding the brain alterations that originate preoperatively.

Altered brain development in CHD is multifactorial. Several risk factors for altered brain growth and brain injury have been identified in the perioperative period. Older postnatal age at surgery (14,16) and longer cardiopulmonary bypass (CPB) duration (14,15) have been associated with reduced brainstem and deep grey matter development (14), as well as new postoperative white matter injury (15,16). In addition, longer postoperative length of pediatric intensive care unit (PICU) stay has been associated with impaired growth of the cortical and deep grey matter, white matter, cerebellum, brainstem (14), and total brain growth (13,14), likely reflecting illness severity and clinical instability (14).

This study aims to use a data-driven Structural Covariance (SC) analysis to (1) assess spatial patterns of coordinated brain development in neonates with CHD after cardiac intervention compared to healthy controls, (2) examine pre- to postoperative changes in coordinated brain development in CHD, and (3) explore the potential effects of perioperative risk factors on these pre- to postoperative changes.

## Methods and Materials

### Ethics and consent

MRI data of healthy neonates were retrieved from the open-access developing Human Connectome Project (dHCP) third data release (17). MRI data of neonates with CHD were acquired as part of The Congenital Heart Disease Imaging Programme (CHIP). Ethical approval was obtained from the National Research Ethics Service West London committee (dHCP: 14/LO/1169; CHIP: 07/H0707/105 and 21/WA/0075) and informed written parental consent was obtained before acquisition of MRI.

### Participants

Neonates with CHD were recruited at birth from St. Thomas’ Hospital, United Kingdom. Neonates with CHD were eligible for inclusion if they had a diagnosis of critical or serious CHD, defined as requiring catheterization or surgery within the first year of life (12,18), were born after 35.00 gestational weeks, and underwent postoperative brain MRI between the postmenstrual age (PMA) of 37.00 and 46.00 weeks. Exclusion criteria comprised known or suspected genetic abnormalities and evidence of major lesions on postoperative brain MRI [arterial ischemic stroke, parenchymal haemorrhage, or >10 foci of white matter injury]. Of 44 eligible neonates, two were excluded due to a diagnosis of 22q11 deletion syndrome and one due to arterial ischemic stroke on postoperative MRI, resulting in a final CHD cohort of 41 neonates. All neonates were previously included in the preoperative CHD cohort reported by Wilson et al. (19).

Neonates were assigned 1 of 3 CHD categories based on hemodynamic impact of the cardiac abnormality using the sequential segmental approach (20), as previously described (11,14,19): abnormal streaming of blood, left-sided cardiac lesions, or right-sided cardiac lesions (see Table 1).

**Table 1.** Demographic details of the cohort. Normally distributed data is presented as mean ± SD, non-normally distributed data is presented as median (IQR), and categorical data is presented as n (%).

|  | CHD<br>(n = 41) | Healthy<br>controls<br>(n = 359) | p |
| --- | --- | --- | --- |
| **Baseline characteristics** |  |  |  |
| Gestational age at birth, median (IQR), *wks* | 38.57<br>(38.00 - 38.86) | 40.14<br>(39.14 - 41.00) | <.001 |
| Gestational age between 35.0 and 36.5, *n (%)* | 2<br>(0.05) |  |  |
| Postmenstrual age at preoperative MRI, median (IQR), *wks* | 39.29<br>(38.71 - 39.86) |  |  |
| Postmenstrual age at postoperative MRI, median (IQR), *wks* | 41.86<br>(40.86 - 42.86) | 41.43<br>(40.14 - 42.86) | 0.172 |
| Male, *n (%)* | 20<br>(48.78) | 185<br>(51.53) | 0.866 |
| Birthweight, median (IQR), *g* | 3140<br>(2870 - 3630) | 3400<br>(3085 - 3728) | 0.009 |
| Birthweight z-score, mean (SD) | -0.11<br>(1.06) | 0.22<br>(0.95) | 0.552 |
| **CHD diagnosis** |  |  |  |
| Abnormal streaming, *n (%)* | 23<br>(56.10) |  |  |
| Transposition of the great arteries, *n (%)* | 20<br>(48.78) |  |  |
| Truncus arteriosus, *n (%)* | 2<br>(4.88) |  |  |
| Total anomalous pulmonary venous drainage, *n (%)* | 1<br>(2.44) |  |  |
| Left-sided lesions, *n (%)* | 13<br>(31.71) |  |  |
| Coarctation of the aorta, *n (%)* | 9<br>(21.95) |  |  |
| Hypoplastic aortic arch, *n (%)* | 2<br>(4.88) |  |  |
| Aortic stenosis, *n (%)* | 2<br>(4.88) |  |  |
| Right-sided lesions, *n (%)* | 5<br>(12.20) |  |  |
| Pulmonary atresia, *n (%)* | 2<br>(4.88) |  |  |
| Pulmonary stenosis, *n (%)* | 2<br>(4.88) |  |  |
| Tetralogy of Fallot, *n (%)* | 1 (2.44) |  |  |
| **Clinical characteristics** |  |  |  |
| Postnatal age at intervention, median (IQR), *wks* | 10<br>(7 - 14) |  |  |
| Cardiopulmonary bypass, *n (%)* | 33<br>(80.49) |  |  |
| Cardiopulmonary bypass time, median (IQR), *min* | 134<br>(104 - 153) |  |  |

MRI data of 359 healthy neonates were retrieved from the developing Human Connectome Project (dHCP) third data release (17), forming the control cohort. Neonates were included if they were born after 37.00 gestational weeks, underwent cerebral MRI between the PMA of 37.00 and 46.00 weeks, had no evidence of major brain lesions on MRI, and did not exhibit severe neurodevelopmental impairments at 18 months of age. Neonates were eligible for inclusion if cognitive and motor scores were no more than 2 standard deviations below the population mean (>70, normative mean 100), assessed using the Bayley-III Scales of Infant and Toddler Development (21).

### Clinical data

Clinical data were extracted from digital patient records, including demographic data, postnatal age at surgery, CPB use and duration, and PICU length of stay. Birthweight was converted to z-scores using the UK90 growth centiles implemented in GrowthCharts v2.0.1. (22).

### MRI acquisition

Neonates were scanned on a 3T Philips Achieva system situated on the neonatal unit at St. Thomas’ Hospital, London, using a 32-channel neonatal head coil and neonatal positioning system (23). The scans included a 5s noise ramp up to prevent a startle response. Pulse oximetry, electrocardiography, respiratory rate, and temperature were monitored throughout scanning. T2-weighted (T2w) multi-slice turbo spin echo scans were acquired in two stacks in sagittal and axial planes (TR =12000ms, TE =156ms, flip angle =90°; slice thickness =1.6 mm; slice overlap =0.8 mm; in-plane resolution: 0.8×0.8 mm; sensitivity encoding factor =2.11/2.58 (axial/sagittal)). T2w volumes were reconstructed using a dedicated algorithm to correct motion and integrate data from both acquired stacks (reconstructed voxel size =0.5 mm^3^) (24,25). T2w images were bias-corrected, brain-extracted and segmented into tissue types (to improve registration), using the dHCP structural pipeline (26).

### Image Processing and Jacobian Determinant Calculation

The pipeline utilized for the extraction and analysis of SC components (SCCs) is summarized in Figure 1. Deformation fields were computed from the non-linear registration of the native T2w images to a template corresponding to the neonate’s PMA at scan from the extended dHCP atlas (17), using symmetric diffeomorphic image registration in Advanced Normalization Tools (27). The warps were concatenated between native T2w, the age-matched template, and a 40-gestational week template from the extended dHCP atlas (17).

**Figure 1.**
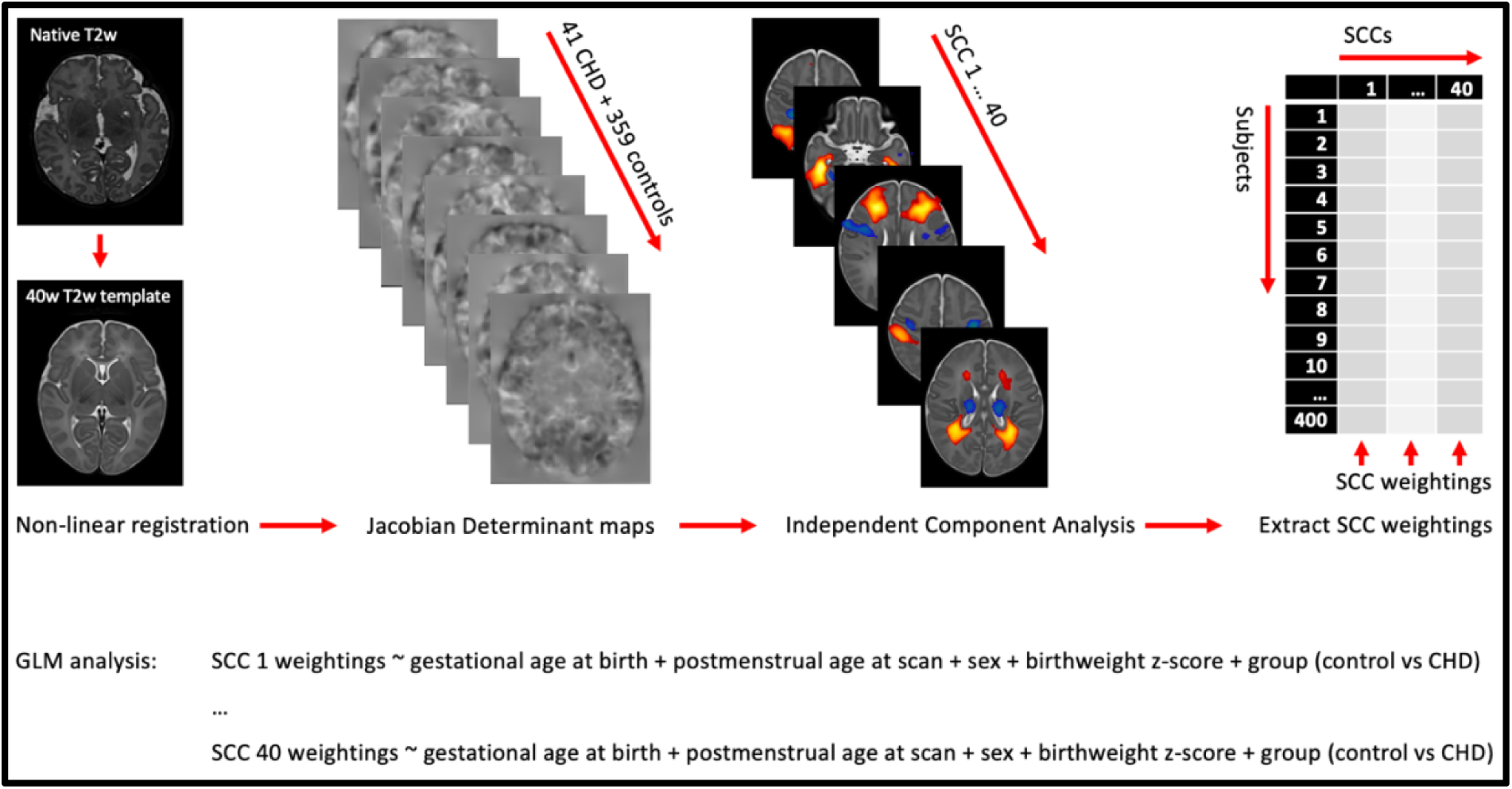
SCC extraction and analysis pipeline. Native T2w images were non-linearly registered to a 40-gestational week T2w template. The warps from registration were used to calculate Jacobian Determinant maps, which were input into an Independent Component Analysis to identify SCCs. SCC weightings were extracted by fitting a GLM to the Jacobian Determinant maps and SCCs. The weightings per SCC were included as a dependent variable in the GLM analysis.

Jacobian determinants were calculated based on the concatenated warps from registration, reflecting the local contraction (higher values) and expansion (lower values) of brain structures on T2w imaging relative to the template (28).

### Independent Component Analysis

The Jacobian determinants of the control group were input into a canonical independent component analysis (CanICA) (29) using the nilearn package in Python (30). This algorithm breaks down the input data into components (i.e., SCCs), effectively separating the mixed signals into independent components of coordinated volume expansion or contraction with non-Gaussian distributions.

We chose 40 as the optimal number of SCCs for our cohort, based on previously reported methodology in SC research (8,31,32), as well as visual assessment of various numbers of SCCs to ensure a balance between robustness and interpretability. Lower numbers of components (i.e., 30 & 35) grouped multiple brain regions together into single components, whereas a higher number of components (i.e., 45) resulted in splitting of bilateral brain regions into separate components. Consistent with previous SC literature (8), the anatomical labels of the SCCs correspond to the positive clusters identified by CanICA (i.e., voxels with values > 0). Additionally, each SCC includes negative clusters (i.e., voxels with values < 0). These positive and negative clusters represent regions that covary inversely (i.e., when one region expands during registration, the other contracts, and vice versa).

### SCC weighting extraction

Weightings were extracted for each SCC by applying the general linear model from FMRIB’s Software Library (33) to the independent component maps and the Jacobian determinant images of healthy controls and neonates with CHD before and after surgery. The resulting SCC weightings reflect the beta values per dataset per SCC, indicative of the strength of relationship between the spatial covariance pattern within that SCC and an individual infant’s Jacobian determinant map.

### Statistical analysis

Rstudio was used for the analyses (v4.2.0) (34). Shapiro Wilk tests were used to assess normality of numerical variables. Data was reported as mean ± SD for normally distributed variables, median (IQR) for non-normally distributed variables, and number (%) for categorical variables. The differences between the CHD group and healthy controls regarding demographic variables were assessed using t-tests for normally distributed numerical variables, Mann-Whitney U tests for non-normally distributed numerical variables, and Chi-square tests for categorical variables.

### SCC differences between CHD and controls

To identify which SCCs differed between neonates with CHD and controls, generalised linear models (GLM) were fit, including SCC weights as the dependent variable and group (CHD vs control) as the independent variable, covarying for gestational age (GA) at birth, PMA at scan, sex, and birthweight z-score.

*Formula: SCC weights ∼ GA at birth + PMA at scan + sex + birthweight z-score + group*

Global Validation of Linear Model Assumptions (GVLMA) was run to determine whether the GLMs violated any assumptions of linearity (i.e., skewness, kurtosis, link function, or heteroscedasticity) (35). For GLMs that violated one or multiple assumptions, model diagnostics were further inspected to assess the extent of the violation (see supplementary Table S1). If assumptions were violated, a robust regression was performed using the same model (36).

### Pre- to postoperative differences in CHD

Linear mixed effect models were used to assess the pre- to postoperative differences in SCC weighting in neonates with CHD, adjusting for GA at birth, PMA at scan, time between scans in days, sex, and birthweight z-score. Multicollinearity was assessed using the variance inflation factor (VIF) and all variables were retained within the model: Scan timepoint (pre- or postoperative) (VIF = 2.63), GA at birth (VIF = 1.70), PMA at scan (VIF = 3.40), time between scans in days (VIF = 1.44), sex (VIF = 1.03), and birthweight z-score (VIF = 1.15). Marginal means adjusted for PMA at scan and time between scans in days were used to estimate mean pre- and postoperative SCC weightings and their difference, with 95% confidence intervals. Residuals of the linear mixed effect models were visually examined using Q-Q plots. Shapiro-Wilk tests were performed for models exhibiting minor deviations from normality, yet no deviations from normality were observed.

### Perioperative risk factors in CHD

Preoperative SCC weightings were subtracted from postoperative SCC weightings for each neonate. Partial Spearman’s rank correlations were then used to investigate associations between pre- to postoperative differences in SCC weightings and perioperative risk factors, including age at surgery, CPB duration, and postoperative PICU length of stay, while adjusting for GA at birth, sex, PMA at postoperative MRI, time between scans in days, and birthweight z-score.

Benjamini–Hochberg False Discovery Rate (FDR) was applied to correct for multiple comparisons. P_FDR_ values < 0.05 were considered significant.

## Results

### Participants

Demographic characteristics of the cohort are presented in Table 1. Neonates with CHD were born at a younger GA (*p* < 0.001) and at a lower birthweight (*p* = 0.009) than healthy controls, however birthweight z-score did not differ between groups. The other demographic variables did not differ significantly between groups.

### SCC differences between CHD and controls

SCCs extracted are shown in Figure 2. Sixteen SCCs were significantly different between neonates with CHD after cardiac surgery and controls (Table 2).

**Figure 2.**
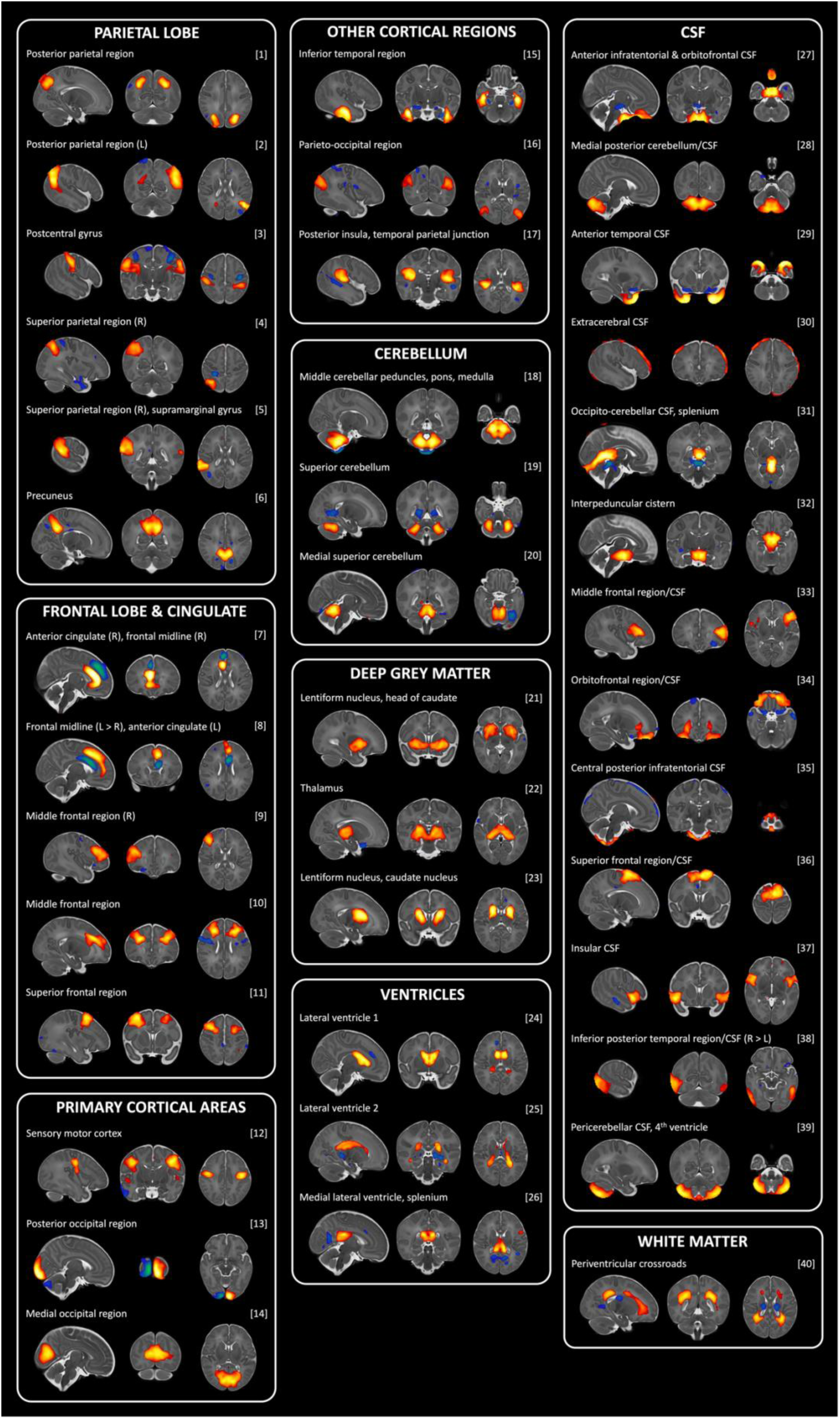
All 40 structural covariance components. Regions within the positive clusters (yellow-red) covary, regions within the negative clusters (blue-green) covary, and positive and negative clusters covary inversely. Anatomical labels correspond to the largest clusters. *Abbreviations:* CSF = cerebrospinal fluid, L = left hemisphere, R = right hemisphere

**Table 2.** Results of GLM or robust regression comparing SCC weightings in neonates with CHD after surgery versus controls, and results of linear mixed-effects models assessing pre- to postoperative changes in SCC weightings in neonates with CHD, adjusted for covariates. Sixteen SCCs differed significantly between CHD and controls, 16 SCCs changed significantly pre- to postoperatively, with 7 SCCs overlapping. *Abbreviations:* CSF = cerebrospinal fluid, P_FDR_ = False Discovery Rate corrected p-value * significant at 0.05 ** significant at 0.05 **^a^** Results of robust regression

|  |  | Postoperative CHD vs controls |  |  | Pre- to postoperative change in CHD |  |  |
| --- | --- | --- | --- | --- | --- | --- | --- |
| SCC | Network label | Beta (95% CI) | t-value | P<sub>FDR</sub> | Beta (95% CI) | F-value | P<sub>FDR</sub> |
| Parietal lobe |  |  |  |  |  |  |  |
| 1 | Posterior parietal region | -3.78 (-10.15 – 2.59) | -1.16 | 0.340 | -1.16 (-7.60 – 5.27) | 11.202 | 0.007** |
| 2 | Posterior parietal region (L) | 0.30 (-0.07 – 0.67)<sup>a</sup> | -1.60<sup>a</sup> | 0.192<sup>a</sup> | -0.70 (-6.53 – 5.12) | 19.035 | 0.001** |
| 3 | Postcentral gyrus | -6.14 (-12.72 – 0.44) | -1.83 | 0.130 | -0.65 (-5.46 – 4.15) | 1.110 | 0.460 |
| 4 | Superior parietal region (R) | 0.16 (-0.21 – 0.53)<sup>a</sup> | 0.84<sup>a</sup> | 0.501<sup>a</sup> | -4.11 (-9.79 – 1.57) | 0.916 | 0.493 |
| 5 | Superior parietal region (R), supramarginal gyrus | -3.52 (-9.46 – 2.42) | -1.16 | 0.340 | -1.25 (-5.23 – 2.73) | 0.057 | 0.879 |
| 6 | Precuneus | -0.05 (-0.43 – 0.34)<sup>a</sup> | -0.24<sup>a</sup> | 0.878<sup>a</sup> | -4.37 (-10.62 – 1.89) | 2.155 | 0.261 |
| Frontal lobe & cingulate |  |  |  |  |  |  |  |
| 7 | Anterior cingulate (R), frontal midline (R) | 0.16 (-0.22 – 0.55)<sup>a</sup> | 0.82<sup>a</sup> | 0.501<sup>a</sup> | -1.77 (-7.96 – 4.42) | 0.816 | 0.513 |
| 8 | Frontal midline (L > R), anterior cingulate (L) | -5.94 (-12.09 – 0.21) | -1.89 | 0.123 | 1.73 (-4.38 – 7.84) | 0.013 | 0.918 |
| 9 | Middle frontal region (R) | 0.58 (0.22 – 0.95)<sup>a</sup> | 3.13<sup>a</sup> | 0.009<sup>a**</sup> | 3.88 (-1.20 – 8.96) | 4.471 | 0.078 |
| 10 | Middle frontal region | -2.25 (-7.53 – 3.03) | -0.84 | 0.520 | -5.78 (-9.70 – -1.86) | 0.011 | 0.918 |
| 11 | Superior frontal region | -4.35 (-10.43 – 1.73) | -1.40 | 0.250 | -3.56 (-9.20 – 2.08) | 0.091 | 0.874 |
| Primary cortical areas |  |  |  |  |  |  |  |
| 12 | Sensory motor cortex | -7.76 (-13.13 – -2.38) | -2.83 | 0.017* | 0.24 (-3.91 – 4.40) | 0.330 | 0.711 |
| 13 | Posterior occipital region | -2.38 (-7.94 – 3.18) | -0.84 | 0.520 | 3.17 (-0.51 – 6.85) | 5.180 | 0.062 |
| Other cortical regions |  |  |  |  |  |  |  |
| 14 | Inferior temporal region | 1.20 (-4.29 – 6.69) | 0.43 | 0.783 | 0.68 (-2.59 – 3.96) | 6.550 | 0.040* |
| 15 | Medial occipital region | -15.59 (-23.57 – -7.61) | -3.83 | 0.002** | -4.15 (-9.58 – 1.27) | 12.981 | 0.004** |
| 16 | Parieto-occipital region | -9.74 (-16.23 – -3.26) | -2.95 | 0.014* | 2.54 (-2.28 – 7.37) | 6.860 | 0.037* |
| 17 | Posterior insula, temporal parietal junction | -0.51 (-6.69 – 5.67) | -0.16 | 0.946 | -0.20 (-4.43 – 4.02) | 15.603 | 0.002* |
| Cerebellum |  |  |  |  |  |  |  |
| 18 | Middle cerebellar peduncles, pons, medulla | 0.62 (0.24 – 1.00)<sup>a</sup> | -3.20<sup>a</sup> | 0.009<sup>a**</sup> | 3.54 (-2.06 – 9.15) | 0.943 | 0.493 |
| 19 | Superior cerebellum | 0.97 (-3.71 – 5.66) | 0.41 | 0.783 | -6.26 (-12.55 – 0.04) | 4.681 | 0.074 |
| 20 | Medial superior cerebellum | 4.43 (-0.20 – 9.06) | 1.87 | 0.123 | 1.99 (-2.82 – 6.80) | 0.508 | 0.640 |
| **Deep grey matter** |  |  |  |  |  |  |  |
| **21** | Lentiform nucleus, head of caudate | -5.22 (-9.20 – 1.24) | -2.57 | 0.028* | -1.34 (-5.38 – 2.70) | 0.447 | 0.655 |
| **22** | Thalamus | -3.50 (-7.01 – 0.01) | -1.96 | 0.114 | 1.67 (-2.64 – 5.97) | 4.811 | 0.072 |
| **23** | Lentiform nucleus, caudate nucleus | -7.22 (-10.72 – -3.71) | -4.04 | 0.002** | 1.20 (-1.92 – 4.32) | 1.109 | 0.460 |
| **Ventricles** |  |  |  |  |  |  |  |
| **24** | Lateral ventricle 1 | 7.99 (2.49 – 13.48) | 2.85 | 0.017* | -1.01 (-6.37 – 4.36) | 35.471 | 0.001** |
| **25** | Lateral ventricle 2 | -0.31 (-0.71 – 0.08)<sup>a</sup> | -1.55<sup>a</sup> | 0.202<sup>a</sup> | -1.76 (-6.33 – 2.80) | 2.509 | 0.220 |
| **26** | Medial lateral ventricle, splenium | -4.78 (-10.25 – 0.69) | -1.71 | 0.159 | -4.54 (-8.42 – -0.66) | 5.969 | 0.048* |
| **CSF** |  |  |  |  |  |  |  |
| **27** | Anterior infratentorial CSF, orbitofrontal CSF | -0.62 (-0.96 – -0.28)<sup>a</sup> | -0.62<sup>a</sup> | 0.005<sup>a**</sup> | 4.05 (1.48 – 6.62) | 92.666 | 0.001** |
| **28** | Medial posterior cerebellum/CSF | 3.20 (-1.28 – 7.69) | 1.40 | 0.250 | 4.16 (-0.70 – 9.01) | 19.589 | 0.001** |
| **29** | Anterior temporal CSF | -10.91 (-18.98 – -2.83) | -2.65 | 0.024* | 3.11 (-3.04 – 9.26) | 9.155 | 0.012* |
| **30** | Extracerebral CSF | 0.44 (0.15 – 0.73)<sup>a</sup> | 3.02<sup>a</sup> | 0.012<sup>a*</sup> | -0.10 (-11.26 – 11.06) | 5.540 | 0.056 |
| **31** | Occipito-cerebellar CSF, splenium | -0.22 (-0.61 – 0.18) | -1.09<sup>a</sup> | 0.383<sup>a</sup> | 7.38 (-0.33 – 15.09) | 35.501 | 0.001** |
| **32** | Interpeduncular cistern | 5.88 (2.46 – 9.29) | 3.37 | 0.008** | 3.97 (-0.90 – 8.84) | 9.264 | 0.012* |
| **33** | Middle frontal region/CSF (L) | -1.83 (-7.11 – 3.46) | -0.68 | 0.604 | 0.07 (-3.13 – 3.26) | 0.060 | 0.879 |
| **34** | Orbitofrontal region/CSF | -0.09 (-5.33 – 5.15) | -0.03 | 0.972 | -1.16 (-4.17 – 1.86) | 23.936 | 0.001** |
| **35** | Central posterior infratentorial CSF | 0.33 (0.07 – 0.58)<sup>a</sup> | 2.53<sup>a</sup> | 0.030<sup>a*</sup> | 10.82 (3.89 – 17.75) | 166.965 | 0.001** |
| **36** | Superior frontal region/CSF | -0.47 (-6.40 – 5.45) | -0.16 | 0.946 | -4.54 (-9.27 – 0.20) | 10.987 | 0.007** |
| **37** | Insular CSF | 10.03 (3.86 – 16.22) | 3.18 | 0.009** | 6.00 (0.43 – 11.56) | 0.107 | 0.874 |
| **38** | Inferior posterior temporal region/CSF (R > L) | -8.42 (-13.56 – -3.29) | -3.21 | 0.009** | 0.41 (-4.15 – 4.97) | 0.022 | 0.918 |
| **39** | Pericerebellar CSF, 4th ventricle | 4.90 (0.11 – 9.69) | 2.00 | 0.108 | 7.79 (4.94 – 10.64) | 0.261 | 0.742 |
| **White matter** |  |  |  |  |  |  |  |
| **40** | Periventricular crossroads | -9.34 (-16.08 – -2.60) | -2.72 | 0.021* | 4.59 (-3.34 – 12.51) | 2.046 | 0.268 |

### Pre- to postoperative differences and perioperative risk factors

The weightings of 16 SCCs were significantly different between the pre- and postoperative MRI (Table 2). The differences in SCC weightings from pre- to postoperative MRI were not significantly associated with age at surgery, CPB duration, or postoperative PICU time (Table 3).

**Table 3.**
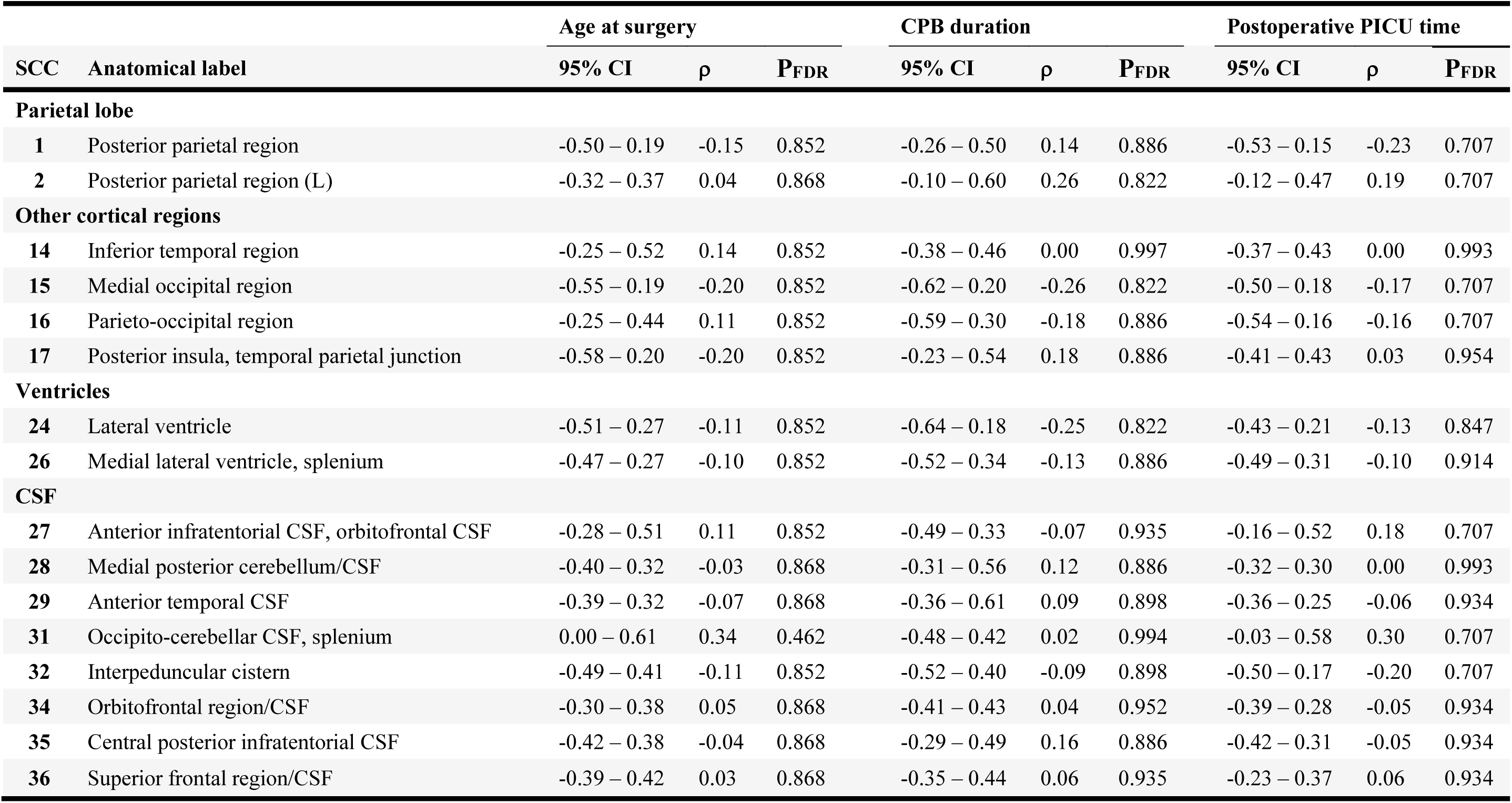
Results of partial correlations between the difference in SCC weightings from pre- to postoperative MRI and perioperative risk factors in neonates with CHD, covarying for gestational age at birth, postmenstrual age at scan, and birthweight z-score. *Abbreviations:* CSF = cerebrospinal fluid, P_FDR_ = False Discovery Rate corrected p-value

## Discussion

This study used a data-driven approach to characterise altered patterns of coordinated brain development in neonates with CHD after cardiac intervention compared to controls and to examine how these patterns change across the perioperative period. Results demonstrated region-specific alterations in coordinated regional brain development in neonates with CHD after surgery compared to controls, and pre-to-postoperative changes within the CHD sample. However, brain changes were unrelated to the perioperative risk factors assessed.

SCCs captured patterns of positive and negative covariance, highlighting the dynamic nature of brain development. Sixteen SCCs showed significant differences between neonates with CHD after cardiac intervention and healthy neonates. The anatomical localisation of these SCCs aligned with previous volumetric studies reporting altered brain development in neonates with CHD in the preoperative (10–12,37) and postoperative (13,14,38) period, including reduced cortical grey matter (10,13,14,37,38), white matter (13,14,37), cerebellar (11,13,14,37,38), brainstem (11,37), and basal ganglia volumes (10,11,37), as well as enlarged CSF spaces (10–12). Additionally, changes identified in the current postoperative analysis overlapped with SCC-derived patterns of altered development observed in a cohort of neonates with CHD scanned before surgery, which partially overlaps with our postoperative sample. These regions comprised the caudate, lentiform, temporal, parietal, occipital, cerebellar, insular, and extracerebral CSF regions (19). Similarly, SCC analysis in fetal CHD revealed altered morphology of extracerebral CSF spaces (8). The overlap of regions across the pre-, and postoperative periods suggests some alterations in CHD likely originate before surgical intervention and persist into the postoperative period.

Neonates with CHD showed significant pre- to postoperative changes in weightings across 16 SCCs. Although SCC analysis does not allow direct inference of regional volumes, these perioperative changes can be interpreted in the context of previous volumetric studies. Cortical (10–12,37), cerebellar (10–12,37), and white matter (11–13,37,39) volumes have been reported to be smaller in neonates with CHD before surgery, while CSF spaces are enlarged (10–12), and ventricular volumes are not different from controls (10–12). In the current study, SCCs encompassing parietal and occipital, lateral ventricle, and temporal and infratentorial CSF regions demonstrated both perioperative change and postoperative differences from controls. It is possible that alterations in these cortical and extracerebral CSF regions originate prior to surgery and become more pronounced into the postoperative period, whereas ventricular morphology may emerge during the perioperative period, consistent with prior reports of postoperative lateral ventricle enlargement in neonates with CHD (40,41). In contrast, SCCs involving frontal, parietal, temporal, cerebellar, and splenium regions, as well as frontal, occipital, and cerebellar CSF regions, showed a perioperative change without a postoperative difference from controls, possibly indicating attenuation of preoperative alterations. The underlying reasons for these patterns remain unclear, however they may relate to the pattern of brain maturation, following a posterior-to-anterior and central-to-peripheral gradient (42–46). Regions that are relatively more mature, such as occipital regions, may be more vulnerable to early disruptions associated with CHD and therefore more likely to demonstrate persistent morphological alterations. In contrast, regions that are relatively less mature, including frontal regions, may show more benefit from improved cardiac physiology following corrective surgery, exhibiting attenuation or a partial catch-up in coordinated development. Overall, these findings highlight heterogeneous, region-specific patterns of altered perioperative brain development in neonates with CHD. Future studies integrating structural covariance and volumetric measures are needed to clarify how perioperative SCC changes correspond to region-specific structural alterations in this population.

The difference between pre- and postoperative SCC weightings was not associated with age at surgery, CPB duration, or postoperative PICU time. Older postnatal age at surgery (14), duration of CPB (14) and length of PICU stay (13,14) have previously been associated with impaired brainstem and basal ganglia development (14), and reduced regional (14) and total (13,14) brain volume in the postoperative period. It is possible that pre-to postoperative changes observed in this study reflect region-specific patterns of altered morphology due to underlying cardiac pathology rather than by perioperative risk factors. Additionally, it was not possible to assess all potential clinical exposures that may influence brain development, and clinical factors not assessed in this study may contribute to these patterns. Finally, although we adjusted for postmenstrual age at MRI, we cannot exclude the possibility that some of the observed changes may reflect typical maturation. Inclusion of longitudinal control cohorts in future studies may help disentangle effects of altered cardiac physiology or perioperative risk factors from maturational processes.

The highlighted regions of altered postoperative brain development in this work have previously been associated with neurodevelopmental outcomes in CHD. Specifically, reduced total and regional brain volumes (13,38), as well as brainstem and basal ganglia volumes (47) in neonates with CHD after cardiac surgery have been associated with poorer language and cognitive development at 1 year (13), poorer motor development in infancy (38) and early childhood (13,38), and below-average IQ at school age (47). SCC analysis in neonates with CHD before cardiac surgery did not reveal any associations between altered brain morphology and neurodevelopmental outcome at 2 years (19). Nonetheless, the overlap between regions showing altered covariance and those previously implicated in cognitive, motor, and language deficits highlights the potential relevance of these patterns for understanding early neurodevelopmental vulnerability in neonates with CHD. Future research including neurodevelopmental assessments is required to investigate the relationship between postoperative region-specific coordinated brain development and neurodevelopmental outcomes.

## Limitations

A limitation of this study is that the distribution of GA differed between the CHD and control group, which might have influenced the results despite the inclusion of GA as a covariate in all analyses. Additionally, the relatively small sample and diagnostic heterogeneity of neonates with CHD (n = 41) may have limited statistical power to detect postoperative differences or perioperative effects. The relatively modest sample size did not to allow for assessment of diagnosis-specific differences in postoperative brain morphology. Further research should include a larger cohort of neonates with a wider variety of CHD diagnoses.

## Conclusion

SCC analysis identified anatomically specific spatial patterns of altered coordinated development in neonates with CHD following cardiac intervention compared to healthy controls, and revealed pre-to-postoperative morphological changes, highlighting heterogeneous, region-specific changes in brain development. These perioperative changes appeared largely independent of commonly reported risk factors, suggesting that these reflect developmental patterns arising from altered cardiac physiology rather than direct effects of surgical exposures. Together, these findings advance our understanding of the dynamic, region-specific nature of perioperative brain development in CHD.

## Data Availability

The data from the dHCP 3rd neonatal release is publicly available (https://nda.nih.gov/edit_collection.html?id=3955). Anonymized derived data from 25 infants with CHD acquired under ethics 21/WA/0075 and infants from dHCP (n = 359), as well as analysis scripts, are available from the corresponding author. We do not have ethical permission for sharing data for 16 infants with CHD acquired under ethics 07/H0707/105.

## Acknowledgements

We would like to thank the families who participated in this study. We thank the staff from the St Thomas’ Neonatal Intensive Care Unit; the Evelina London Children’s Hospital Fetal and Paediatric Cardiology Departments; the Evelina London Paediatric Intensive Care Unit and the Centre for the Developing Brain at King’s College London; and the Evelina Newborn Imaging Centre.

