## Supplementary Material for "Structural Covariance Analysis of Altered Brain Development in Neonates with Congenital Heart Disease After Surgery"

**GLM assumption check**

In addition to the GLM's, GVLMA was run to check whether assumptions of linearity were violated. If GVLMA indicated violation of certain assumptions of linearity, additional tests were run to check whether the violations were truly present. We encountered violations of skewness and kurtosis, for which we ran a robust regression instead. As for the GLM's where GVLMA indicated violations, yet the additional tests did not, GLM results were reported. The violations we encountered and how they were dealt with is summarized in Supplementary Table S1.

| SCC | GVLMA violation | Additional tests | Analysis |
|---|---|---|---|
| [2] Posterior parietal region (L) | Skewness | *Assumption violated*<br>Residuals plots: indicated slight skewness<br>Q-Q plot: indicated slight skewness<br>Shapiro-Wilk test significant ($p$ = 0.042) | Robust regression |
| [4] Superior parietal region (R) | Skewness | *Assumption violated*<br>Residuals plots: indicated skewness<br>Q-Q plot: indicated skewness<br>Shapiro-Wilk test significant ($p$ < 0.001) | Robust regression |
| [6] Precuneus | Skewness + Link function | *Assumption violated*<br>Residuals plots: indicated skewness<br>Q-Q plot: indicated skewness<br>Shapiro-Wilk test significant ($p$ = 0.004)<br>Residuals vs Fitted values plot: indicated violation<br>Observed vs Predicted values plot: indicated violation | Robust regression |
| [7] Anterior cingulate (R), frontal midline (R) | Skewness | *Assumption violated*<br>Residuals plots: indicated skewness<br>Q-Q plot: indicated skewness<br>Shapiro-Wilk test significant ($p$ < 0.001) | Robust regression |
| [9] Middle frontal region (R) | Skewness + Kurtosis | *Assumption violated*<br>Residuals plots: indicated skewness<br>Q-Q plot: indicated skewness<br>Shapiro-Wilk test significant ($p$ = 0.013) | Robust regression |
| [11] Superior frontal region | Link function | *Assumption met*<br>Residuals vs Fitted values plot: did not indicate violation<br>Observed vs Predicted values plot: did not indicate violation | GLM |
| [18] Middle cerebellar peduncles, pons, medulla | Skewness | *Assumption violated*<br>Residuals plots: indicated slight skewness<br>Q-Q plot: indicated skewness<br>Shapiro-Wilk test significant ($p$ = 0.014) | Robust regression |
| [25] Lateral ventricle 2 | Skewness | *Assumption violated*<br>Residuals plots: indicated skewness<br>Q-Q plot: indicated skewness<br>Shapiro-Wilk test significant ($p$ < 0.001) | Robust regression |
| [27] Anterior infratentorial CSF, orbitofrontal CSF | Link function | *Assumption violated*<br>Residuals vs Fitted values plot: indicated violation<br>Observed vs Predicted values plot: indicated violation | Robust regression |

| | | | |
|---|---|---|---|
| [30] Extracerebral CSF | Skewness + Link function | *Assumption violated*<br>Residuals plots: indicated slight skewness<br>Q-Q plot: did not indicate skewness<br>Shapiro-Wilk test not significant ($p = 0.139$)<br>Residuals vs Fitted values plot: indicated violation<br>Observed vs Predicted values plot: indicated violation | Robust regression |
| [31] Occipito-cerebellar CSF, splenium | Kurtosis | *Assumption violated*<br>Residuals plots: indicated skewness<br>Q-Q plot: indicated skewness<br>Shapiro-Wilk test significant ($p < 0.001$) | Robust regression |
| [32] Interpeduncular cistern | Link function | *Assumption met*<br>Residuals vs Fitted values plot: did not indicate violation<br>Observed vs Predicted values plot: did not indicate violation | GLM |
| [35] Central posterior infratentorial CSF | Kurtosis + Link function | *Assumption violated*<br>Residuals plots: did not indicate skewness<br>Q-Q plot: indicated slight skewness<br>Shapiro-Wilk test significant ($p = 0.03$)<br>Residuals vs Fitted values plot: indicated violation<br>Observed vs Predicted values plot: indicated violation | Robust regression |

**Supplementary Table S1.** Summary of assumptions check for GLM models
